# Lower limb joint Loading during high-impact activities: implication for bone health

**DOI:** 10.1101/2024.03.11.24303795

**Authors:** Zainab Altai, Claude Fiifi Hayford, Andrew Phillips, Jason Moran, Xiaojun Zhai, Bernard X.W. Liew

## Abstract

Osteoporosis, a significant concern among the elderly, results in low-trauma fractures affecting millions globally. Despite the inclusion of physical activities in strategies to mitigate osteoporosis-related fractures, the optimal exercises for bone health remain uncertain. Determining exercises that enhance bone mass requires an understanding of loading on lower limb joints. This study investigates hip, knee, and ankle joint loading during walking, running, jumping, and hopping exercises, assessing impacts at various intensities (maximal, medium, and minimum efforts).

A total of 37 healthy, active participants were recruited, with a mean (SD) age of 40.3 (13.1) years, height of 1.7 (0.08) m, and mass of 68.4 (11.7) kg. Motion capture data were collected for each participant while performing six different exercises: a self-selected level of walking, running, counter-movement jump, squat jump, unilateral hopping, and bilateral hopping. A lower body musculoskeletal model was developed for each participant in OpenSim. The static optimization method was used to calculate muscle forces and hip joint contact forces.

The study reveals that running and hopping induce increased joint contact forces compared to walking, with increments of 83% and 21%, respectively, at the hip; 134% and 94%, respectively, at the knee; and 94% and 77%, respectively, at the ankle. Jump exercises exhibit less hip and ankle loading compared to walking, with reductions of 36% and 19%, respectively. Joint loading varies across exercises and intensities, running faster increases forces on all joints, especially the hip. Sprinting raises hip forces but lowers forces on the knee and ankle. Higher jumps intensify forces on the hip, knee, and ankle, whereas hopping faster reduces forces on all joints.

The study emphasizes the site-specific impact of exercises on lower limb joint loadings, highlighting the potential of running and hopping for bone formation compared to jumping alone. These findings offer insights for optimizing exercise routines to improve bone health, with potential implications for risk prevention, rehabilitation, and prosthetic development.

## 1. Introduction

Osteoporosis poses a significant health challenge, particularly among the elderly population. One of the main consequences of osteoporosis is law trauma fractures. Approximately 137 million women and 21 million men worldwide are estimated to face an increased risk of osteoporotic fractures, causing increasingly important public health concerns.^1^ Current strategies aimed to optimizing bone mass to help in reducing the risk of osteoporosis-related fractures include physical activities.^2,3^ It has been reported that normal walking^4^ is not associated with bone mineral density (BMD) changes in the femoral neck whereas jogging combined with walking^5^, running, and jumping^6–8^ were the most effective in improving bone density and other parameters of bone health. Nevertheless, the optimal type and intensity of exercises that enhance bone mass are still largely unknown.^9,10^ That is due to the knowledge gap on how different loading paradigms best protect/enhance bone density and strength.^2,9^

The relationship between physical activities, exercises, and bone health is explained by the “mechanostat” theory^11,12^, which states that bone adapts its microstructure based on the induced mechanical loadings. These loadings are represented by external loading, including ground reaction forces (GRF) and joint moments, and internal loading, including joint contact forces (JCF) and muscle forces. When the imposed force on the bone exceeds a particular threshold, bone formation occurs in favor of bone resorption. According to a meta-analysis by Kistler-Fischbacher et al, a high GRF, of at least two times the body weight, is required to stimulate an osteogenic effect.^13^ However, studies have shown that GRF metrics can mislead our understanding of loading on internal structures.^14^ To allow prediction of the osteogenic effect of different exercises, quantitative data are required on calculated forces acting on the lower limb joints. These forces generate stresses and strains in the bones by which local adaptation of bone microstructure occurs. Recent studies reported that JCF at the hip is strongly related to the strain distribution in the proximal part of the femur during walking^15^, running^16,17^ and jumping.^17^ However, while there are many benefits to high-impact exercise, it may carry potential risks. High-impact movements may impose higher stress than desired on the joints, increasing the risk of both acute and overuse injuries. Accordingly, the potential to relate joint contact forces at the hip, knee, and ankle during different exercises to stresses and strain levels that trigger osteogenesis could be a major advancement toward optimizing training programs that induce appropriate force levels to stimulate bone formation while avoiding injuries.

The “gold standard” method for in vivo measurement of joint contact forces includes using instrumented implants.^18^ However, this method is limited by a small number of subjects in addition to the altered anatomy and physiology of the joint region due to surgery. On the other hand, musculoskeletal modeling based on 3D motion capture data offers advanced tools to predict joint forces comparable to experimental measurements during various physical activities.^19^ Several previous studies have employed musculoskeletal modeling to investigate the effect of different speeds of walking and running on ground reaction forces^20^ and contact forces of various lower limb joints.^21–23^ To the authors’ knowledge, no research on such predictions during different jumping and hopping movements is available. In studies in which exercise intensity is investigated, most focus predominantly on the effect on a single lower limb joint (hip ^16,17,24^, knee ^25,26^, and ankle ^26^) and did not investigate the combined effect of movement on other joints. Altering loading on one lower limb joint may be associated with changes in the loading of the other joints.^27^ One recent study reported increased contact force of three lower limb joints when increasing the speed of gait, but more interestingly, a significant increase was found in the predicted joint contact force at the knee compared to the hip and ankle ^28^. Unfortunately, only walking was investigated.^28^ Niu et al reported that increasing dropping height significantly increased compressive forces of the hip, knee, and ankle joints.^29^ But more importantly, when subjects landed from low (32 cm) and medium heights (52 cm), from the ankle to the knee and the hip joints, the peak joint forces in the vertical direction declined.^29^ However, dropping from a height may produce a different force pattern than jumping from the ground and landing. In terms of the effect of various exercise types, Pellikaan et al found that fast walking, running, and unilateral hopping induced significantly higher hip joint contact forces than walking at 4 km/h.^17^ They advise that hopping needs to be considered carefully, especially in the elderly population. However, no information was reported on knee and ankle joints. As aging is predictive of the development of knee osteoarthritis, a condition often associated with the potential worsening of other existing health issues, a comprehensive examination of all three joints becomes pivotal.

The present study aims to explore alterations in hip, knee, and ankle joint loading across various exercise types (walking, running, countermovement jump, squat jump, unilateral hopping, and bilateral hopping) and intensities (low, moderate, and maximum levels). This investigation seeks to address two primary questions: firstly, which exercises elicit higher loading at the hip, knee, and ankle joints compared to walking at normal speed, and secondly, how exercise intensity influences loading at these three joints. By investigating the type and intensity of exercise, that optimally stimulates bone remodeling without compromising joint integrity, this study seeks to contribute valuable insights to the development of targeted exercise interventions for individuals at risk of osteoporosis-related issues.

## 2. Material and methods

### 2.1. Dataset

A total of 40 healthy participants were recruited for the present study, which occurred in the motion laboratory at the University of Essex, UK. Eligible participants have to have no lower limb joint replacement, have not been diagnosed with any serious lower limb injuries within the last year, and are within the age range of 18 to 70 years old. The final sample includes 20 males and 20 females (age = 40.3 ± 13.1 years; height = 1.7 ± 0.08 m; mass = 68.4 ± 11.7 kg). Ethical approval was obtained from the University of Essex Faculty of Science & Health Ethics Subcommittee (ETH2021-1155). A written consent form was obtained from all participants before participating.

### 2.2. Experimental Setup and Protocol

Thirty-eight retro-reflective markers were attached to the lower extremity for each participant. Twenty-two individual markers were attached to the following anatomical landmarks: left and right superior iliac spines, anterior superior iliac spines and posterior superior iliac spines, medial and lateral femoral condyles, medial and lateral malleoli. On the shoe, rearfoot markers were attached to the calcaneus’s lateral and posterior aspects, while forefoot markers were attached to the first and fifth metatarsals. Furthermore, tracking clusters consisting of 4 markers were attached to the distal lateral aspect of the thigh and the shank (Fig. 1). Five electromyography (EMG) sensors (Norixon, AZ., USA, 2 kHz) were attached unilaterally to the dominant side of each participant targeting five different muscles: gluteus maximus, gluteus medius, rectus femoris, biceps femoris, and soleus. Surface EMG signals were used later to validate the musculoskeletal model’s muscle force predictions. Marker trajectories were recorded using sixteen 3D motion capture cameras (Vicon. Ltd., Oxford, UK, 200Hz). Ground reaction forces were collected using two-floor force plates (Kistler, Winterthur, Switzerland, 2 kHz).

**Fig. 1.**
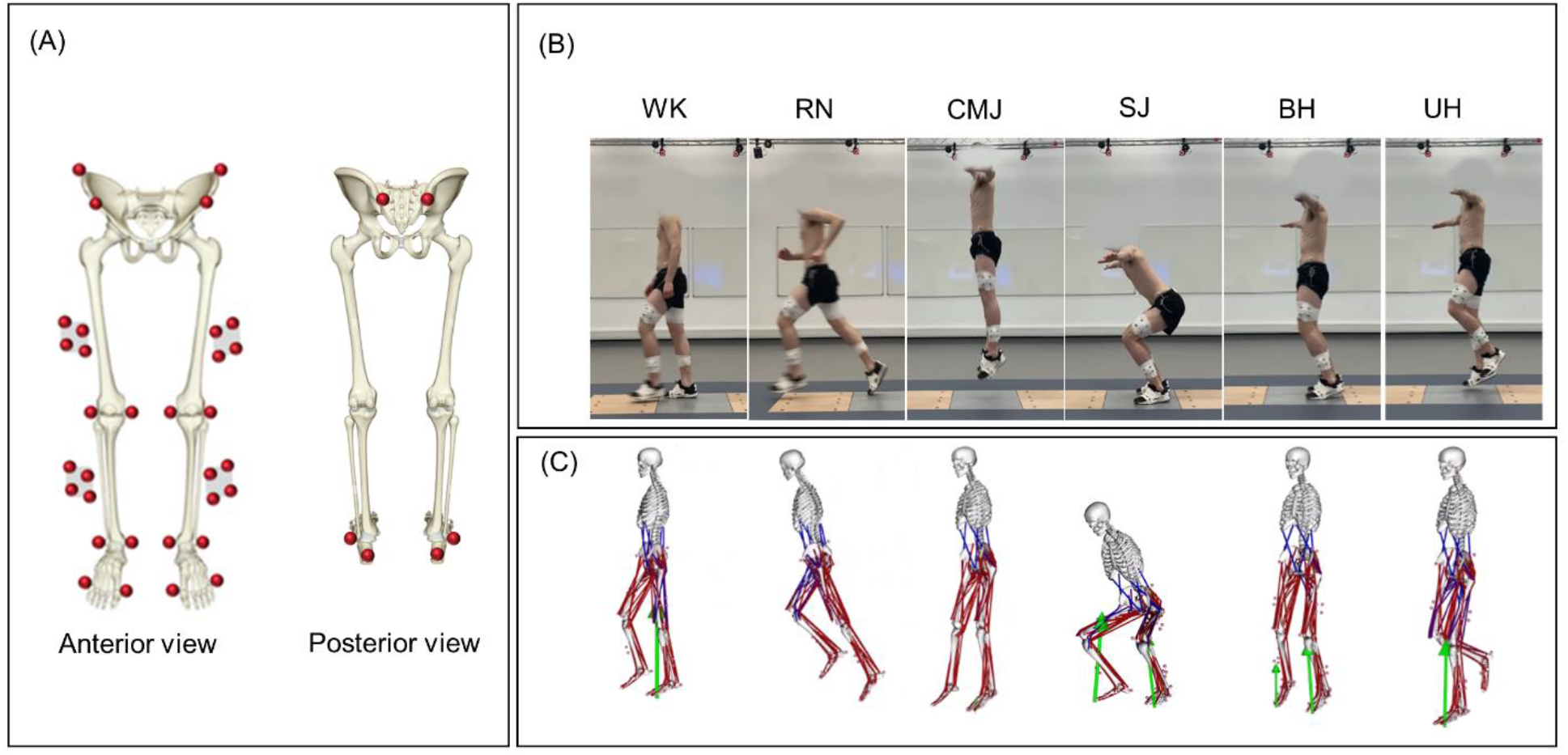
Marker set for motion capture data and Musculoskeletal models. Twenty-two individual markers and four cluster markers were placed on the bone anatomical landmarks of the lower body (A). Six different exercises; walking, running, countermovement jump, squat jump, unilateral hopping, and bilateral hopping, were performed by each participant (B). Scaled Musculoskeletal models were developed in OpenSim representing each performed exercise (C).

Participants performed six different exercises: walking (WK), running (RN), countermovement jump (CMJ), squat jump (SJ), unilateral hopping (UH), and bilateral hopping (BH). Participants were asked to perform each exercise (apart from walking) at three different self-reported intensity levels: maximum effort, medium effort, and minimum effort as follows:

- Running, maximum effort (Fast) was achieved by instructing participants to sprint at their highest attainable speed; minimum effort (Natural) was achieved by asking the participants to maintain their typical jogging pace; while medium effort (Moderate) was determined collaboratively by the participants and the instructor, taking into consideration the parameters of maximum and minimum effort levels.
- Squat jump and counter movement, maximum effort (Max) was achieved by instructing participants to jump at their highest attainable height; while minimum effort (Min) was achieved by asking the participants to perform minimal jump; and again, medium effort (Med) was determined collaboratively by the participants and the instructor, taking into consideration the parameters of maximum and minimum effort levels.
- Unilateral hopping and Bilateral hopping, participants were instructed to hop for 10 seconds to beat three distinct frequencies (3.0 Hz, 2.6 Hz, and 2.2 Hz, representing maximum effort (Max), medium effort (Med), and minimum effort (Min), respectively) ^30^. Beats were played for the participants in advance and were asked to practice until they achieved synchronization.
- Walking trials were collected at a self-selected speed of normal walking.

Participants were allowed to warm up for two minutes before recording the actual trials. Three successful trials (whole foot in contact within the force plate area) were collected for each intensity levels of each exercise.

### 2.3. Musculoskeletal modelling

A generic musculoskeletal model (gait2392) developed by Delp et al^31^ was modified by removing the torso and associated muscles. The modified lower extremity model consisted of 13 body segments, 18 degrees of freedom (DOF), and 86 Hill-type musculotendon actuators. The hip was modelled as a ball and socket joint (3 DOF), while the knee was modeled as a sliding hinge joint (1 DOF), and the ankle and subtalar as revolute joints (1 DOF). Each model was scaled to match the subject’s anthropometric characteristics based on marker data of anatomical landmarks at the hip, knee and ankle during a static trial. The maximal isometric force of each muscle was scaled by the mass of the subject divided by the mass of the generic model raised to the power 2/3. The maximal isometric force was increased by a factor of 3 for successful simulation for some tasks.^32^ A typical Opensim^33^ simulation pipeline was followed to estimate joint angles and moments using inverse kinematics and inverse dynamics, respectively. Static optimization was used to estimate muscle forces by minimizing the sum of squared muscle activations. Using joint reaction analysis,^34^ hip, knee and ankle contact forces (JCFhip, JCFknee, and JCFankle, respectively) were calculated for left and right side. However, data analysis for the current study focused on the participants’ dominant side, determined by asking them which foot they used to kick a ball.

### 2.4. Trial and Data processing

Trials were segmented based on trial type as described in Table 1. Time points used for trial segmentation were defined using Visual 3D (C-motion Inc., Germantown, MD, USA). All trials were then time normalized to 101 time points. Ground reaction forces and joint contact forces were also normalized by the body weight of the participant (BW). Averaging processes were conducted at two levels. A first averaging process resulted in an ensemble average curve across all participants for every exercise intensity of every exercise type. For that, force curves were averaged for the repetitive trials (three trials per exercise per intensity) for each participant across the 101 time points. Then the averaged curves were used to find the mean curves of the vertical ground reaction force (vGRF) and the resultant joint contact forces (JCFhip, JCFknee, and JCFankle) across all participants (supplementary material, Fig. 1, Fig. 2, Fig. 3, respectively). The second averaging process resulted in the mean of the peak value of forces across all participants. Peak values were identified in the averaged curved of each participant then the means of the peaks were calculated for each of exercise intensity across all participants.

**Table 1.** Time points used to segment trials for each of the five tested exercises.

| Exercise type | Time point used for trial segmentation |  |
| --- | --- | --- |
|  | From | To |
| WK | Heel strike | Toe off |
| RN | Heel strike | Toe off |
| BH | Foot on the force plate* | Foot off force plate* |
| UH | Foot on the force plate | Foot off force plate |
| CMJ | Initial stand (just before take-off) | Lowest position of the pelvis after landing |
| SJ | Lowest pelvis position during the squat position (just before take-off) | Lowest position of the pelvis after landing |

\* Dominant side
WK: walking, RN: running, CMJ: countermovement jump, SQ: squat jump, UH: unilateral hopping, and BH: bilateral hopping.

**Fig. 2.**
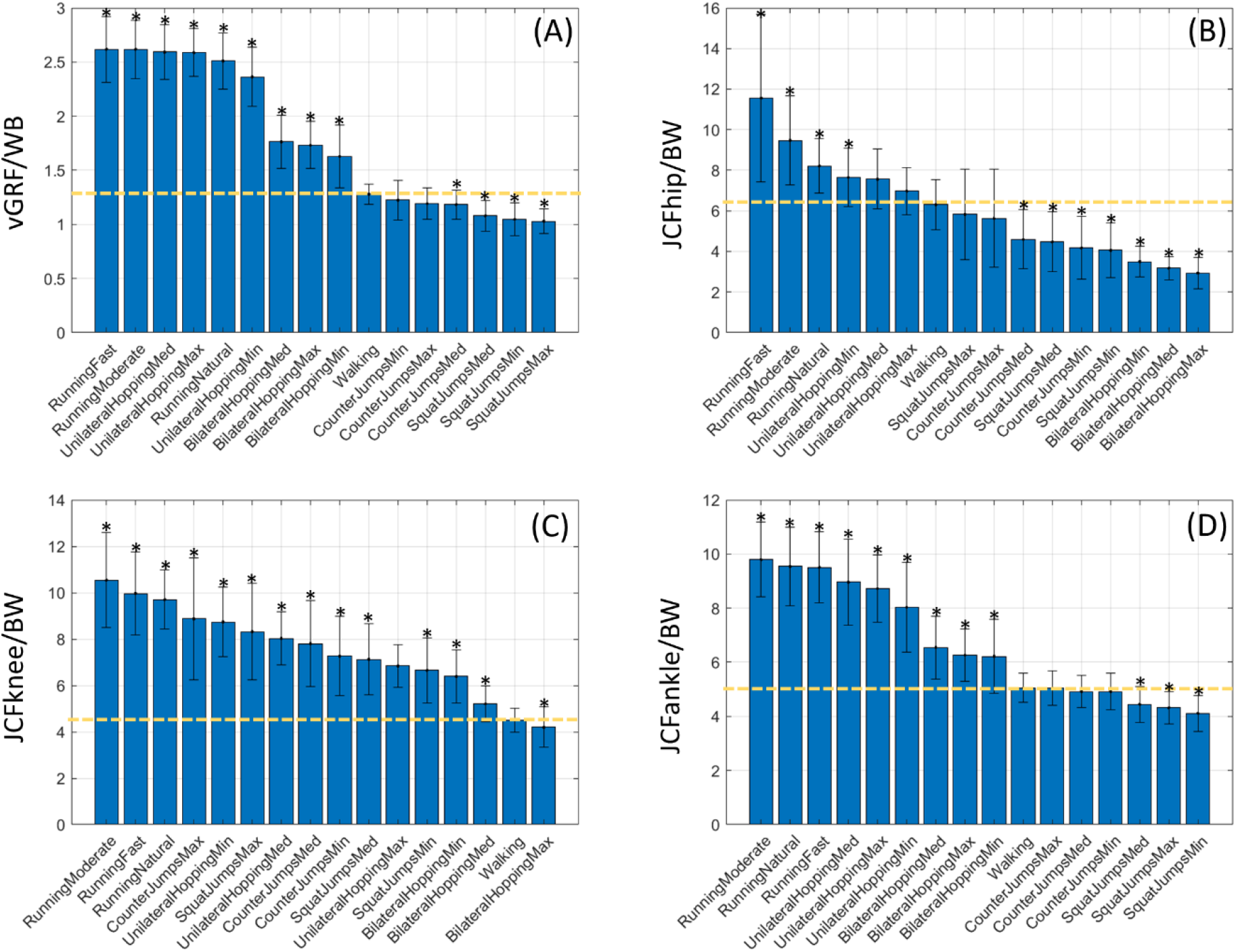
Average peak vertical Ground reaction force (A), resultant joint contact forces of the hip (B), knee Cc), and ankle (D) expressed in body weight of each participant (BW) ranked from left to right for the highest to the lowest predicted force. Asterisks denote the exercises with significantly different (*p < 0.05) peak force compared to walking (highest peak of Peak1 and Peak2) indicated by the horizontal line.

**Fig. 3.**
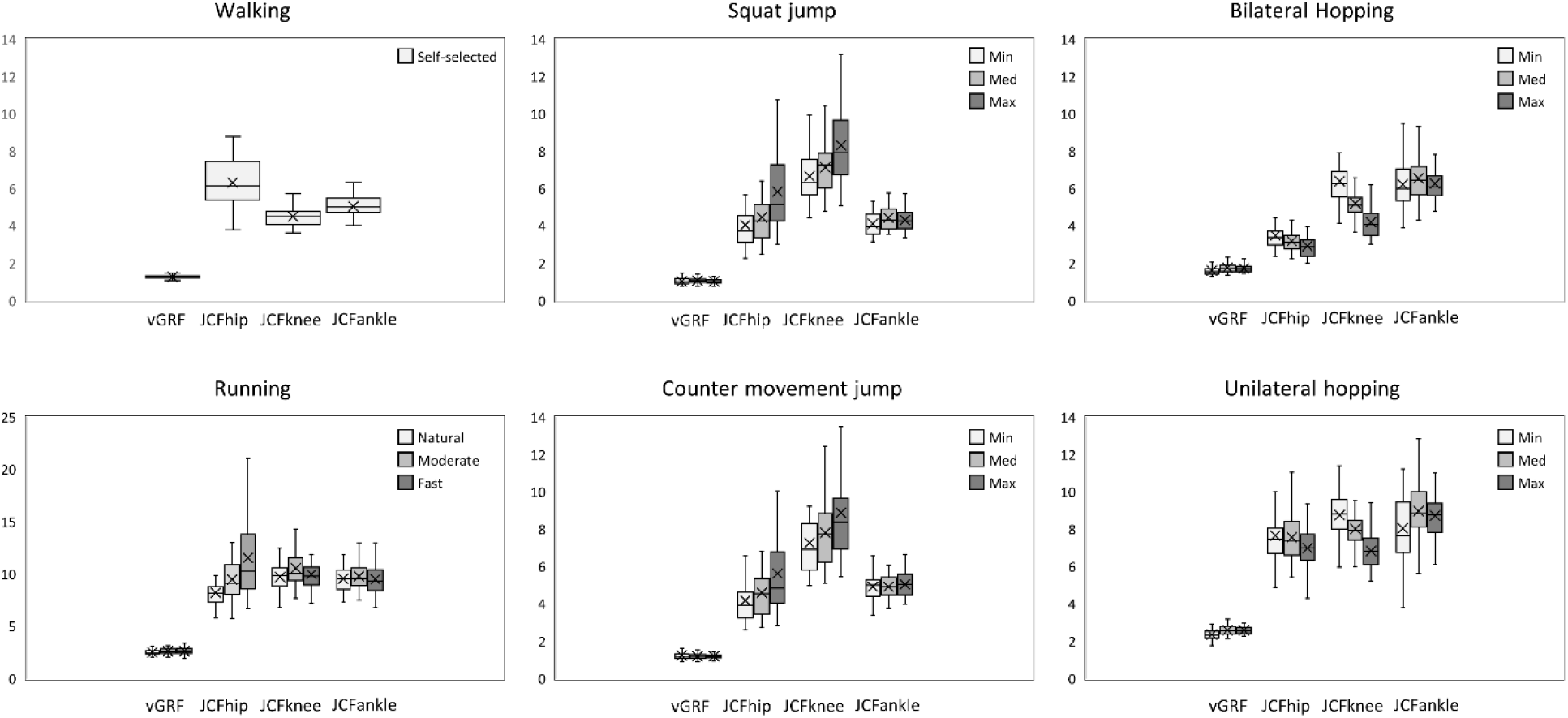
Distribution of peak vGRF and peak JCFs during walking, running, squat jump, counter movement jump, unilateral hopping, and bilateral hopping and at three different exercise intensities. vGRF ground reaction force; JCF contact force at hip, knee, and ankle.

### 2.5. Statistical Analysis

A repeated measures ANOVA was performed on the peaks of the ground reaction forces and joint contact forces of all participants to test the global effect of exercise type and exercise intensity compared to walking using the General Linear Model in SPSS (Chicago, USA). The dependent variable was JCF of each of the hip, knee, ankle, and vGRF, whilst the independent variable was the various exercise types and intensities. Where significance was found (significance level α = 0.05), Bonferroni post hoc test was conducted to quantify pairwise differences.

## 3. Results

The means and standard deviations of the peak vertical ground reaction force, and joint contact forces of the hip, knee, and ankle across all exercise types and intensities are indicated in Table 2.

**Table 2.** Mean and SD of the peak vertical ground reaction force and joint contact forces of the hip, knee and ankle predicted by the musculoskeletal models for the six tested exercises.

| Exercise | Mean±SD peak ground reaction forces and joint contact forces normalised by the body weight |  |  |  | Mean±SD Speed (m/sec) | Mean±SD Jump height (m) | Mean±SD Stance duration (sec) |
| --- | --- | --- | --- | --- | --- | --- | --- |
|  | vGRF | JCFhip | JCFknee | JCFankle |  |  |  |
| Walking | 1.28±0.09 | 6.31±1.23 | 4.50±0.50 | 5.05±0.53 | 1.59±0.41 | - | - |
| Running Natural | 2.51±0.26 | 8.21±1.32 | 9.70±1.26 | 9.54±1.43 | 2.98±0.61 | - | - |
| Running Moderate | 2.62±0.27 | 9.47±2.17 | 10.54±2.02 | 9.79±1.36 | 4.25±0.59 | - | - |
| Running Fast | 2.62±0.30 | 11.56±4.08 | 9.97±1.77 | 9.51±1.30 | 5.26±0.83 | - | - |
| Squat Jumps Min | 1.05±0.15 | 4.06±1.33 | 6.65±1.38 | 4.11±0.66 | - | 0.21±0.06 | - |
| Squat Jumps Med | 1.08±0.14 | 4.47±1.45 | 7.13±1.53 | 4.44±0.65 | - | 0.26±0.08 | - |
| Squat Jumps Max | 1.03±0.11 | 5.83±2.21 | 8.32±2.05 | 4.32±0.59 | - | 0.32±0.09 | - |
| Counter Jumps Min | 1.22±0.18 | 4.18±1.52 | 7.26±1.69 | 4.92±0.67 | - | 0.23±0.05 | - |
| Counter Jumps Med | 1.18±0.13 | 4.59±1.44 | 7.80±1.82 | 4.92±0.58 | - | 0.28±0.07 | - |
| Counter Jumps Max | 1.19±0.14 | 5.63±2.38 | 8.88±2.59 | 5.04±0.64 | - | 0.33±0.08 | - |
| Unilateral Hopping Min | 2.36±0.27 | 7.65±1.43 | 8.74±1.48 | 8.04±1.64 | - | - | 0.31±0.05 |
| Unilateral Hopping Med | 2.59±0.25 | 7.56±1.46 | 8.02±1.13 | 8.96±1.58 | - | - | 0.28±0.06 |
| Unilateral Hopping Max | 2.59±0.22 | 6.98±1.14 | 6.84±0.92 | 8.72±1.23 | - | - | 0.25±0.06 |
| Bilateral Hopping Min | 1.63±0.29 | 3.50±0.74 | 6.39±1.14 | 6.22±1.35 | - | - | 0.25±0.04 |
| Bilateral Hopping Med | 1.76±0.24 | 3.18±0.57 | 5.21±0.76 | 6.55±1.15 | - | - | 0.21±0.03 |
| Bilateral Hopping Max | 1.73±0.22 | 2.93±0.76 | 4.20±0.86 | 6.27±0.96 | - | - | 0.19±0.02 |

vGRF: Vertical ground reaction force
JCFhip: Hip joint contact force
JCFknee: Knee joint contact force
JCFankle: Ankle joint contact force
SD: standard deviation

### 3.1. Exercise type

All tested exercises were ranked based on the average peak force for vertical ground reaction force (Fig. 2 A), hip joint contact force (Fig. 2 B), knee joint contact force (Fig. 2 C), and ankle joint contact force (Fig. 2 D). Exercises with a significant difference (p < .05) compared to walking with self-selected speed (highest peak of Peak1 and Peak2) were marked with an asterisk. The estimates, lower/upper limits, and p-values as well as the results from the ANOVA were reported in the supplementary material (Table 1).

#### 3.1.1 Vertical ground reaction force

Running, unilateral hopping, and bilateral hopping imposed higher vertical ground reaction force compared to walking by up to 105%, 103%, and 38% respectively. While both countermovement jump and squat jump were lower than walking by 8%, and 20% respectively.

#### 3.1.2 Joint contact forces

Running and unilateral hopping imposed higher forces on the hip joint compared to walking, with increases of up to 83% and 21%, respectively. Conversely, forces on the hip joint decreased by 53%, 36%, and 34% during bilateral hopping, squat jumps, and counter-movement jumps, respectively.

All exercises resulted in higher knee joint contact forces compared to walking. Running induced the highest force, increasing by up to 134%, followed by counter movement jumps at 97%, unilateral hopping at 94%, squat jumps at 85%, with the lowest increase during bilateral hopping at 42% higher than walking.

Similar to the hip joint, the ankle joint experienced increased loading during running (94%), unilateral hopping (77%), and bilateral hopping (30%) compared to walking. Conversely, both squat jumps and counter-movement jumps imposed lower forces on the ankle joint than walking, with reductions of 19% and 3%, respectively.

### 3.2. Exercise Intensity

Overall, the impact of exercise intensity on the vertical ground reaction force was barely noticeable across all exercises with only a minor increase observed when hopping at a higher frequency. A clear effect was observed on joint contact forces (Fig. 3). Increasing running speed from jogging to a moderate speed resulted in increased forces across all joints. However, sprinting at the highest capacity exhibited a considerably higher force at the hip joint combined with decreased forces on the knee and ankle joints. Increased vertical jump amplitude caused an increase in forces experienced at the hip, knee, and ankle joints. Conversely, increased hopping frequency results in a reduction in forces exerted across all joints.

## 4. Discussion

The current study explores the effect of high-impact exercises at different intensity levels on hip, knee, and ankle joint loadings. The aim was to evaluate the types and intensity of exercises that may either trigger or support bone tissue formation. Running and hopping at all tested intensities lead to significantly higher joint contact forces in the three joints compared to normal walking speed, potentially stimulating bone remodeling. Hip joint load was higher than the knee and ankle during walking, while knee joint load was higher during running and jumping, and ankle load was higher during hopping.

The range of joint contact forces of the hip, knee, and ankle joints predicted by the current study is comparable to previous studies.^12,16,17,22,35–37^ Prior simulation studies reported hip joint forces during slow to fast runs to be 7.5-10.0 BW ^16^ with speed range (0.8 to 3.6 m/sec), compared to our predicted 8.2-11.6 BW. Knee joint forces were reported around 7.8-12.0 BW at 4.36 m/sec ^12,22^, while our prediction was 9.7-10.5 BW. Ankle joint forces ranged from 11.9 BW to 13 BW at 4.36 m/sec ^22,35^ compared to our prediction of around 10.0 BW. Some of our predicted forces are slightly higher, one reason possibly due to our cohort comprising healthy, active individuals who exercise regularly at least three times a week, as indicated by a questionnaire administered before data collection. This is confirmed by the higher running speed range of the current study cohort (2.98 to 5.25 m/sec) compared to the above-mentioned studies. Another reason could be related to the use of static optimization technique to estimate muscle forces in the musculoskeletal models of the present study. Static optimization methods were reported to likely overestimate lower-limb joint contact forces during vigorous gait tasks.^22^ However, static optimization solutions were also reported to practically equivalent to dynamic solution^38^. In terms of hopping and jumping, despite the limited available information, our results align with the existing data. Pellikaan et al. reported a hip joint contact force range of 6.0-7.57 BW during self-selected unilateral hopping at what pace^17^. The study included post-menopausal elderly women, potentially leading to lower reported values compared to our study. While during maximal countermovement jump performed by athletic males, joint contact forces were reported to be 5.5-8.4 BW, 6.9-9.0 BW, and 8.9-10.0 BW at 0.38 m jump height^39^ compared to our predictions (4.2-5.6 BW, 7.3-8.9 BW and 4.9-5.0 BW at jump height range of 0.23 to 0.33 m) for hip, knee, and ankle joints, respectively. Knee joint contact force during squat jumps was reported to be an average of 7.07 BW^37^ compared to the current study prediction (6.65 BW-8.32BW at jump height range of 0.21 to 0.32 m).

The current study assumes normal walking, which is a low-impact daily activity, as the baseline to judge which exercise is seen as a more potent stimulus for promoting beneficial changes in bone structure and density^4,17,40^ Running and hopping involves brief periods of weightlessness followed by forceful ground impacts and joint loading, inducing a more significant skeletal response. These results are partly in agreement with several clinical trials and simulation studies. Fast-walking intervention program^5,41,42^, running^17^, and hopping^17,43^ were reported to preserve femoral neck BMD in an elderly population. Interestingly, our results suggest no, or even a negative, effect of countermovement jump and squat jump exercises, even when jumps are performed with the maximal effort, in particular at the hip and ankle joint regions at which loading was found to be less than walking. Therefore, jump exercises alone seem to be unsuitable to increase or maintain the BMD. Despite jumping strength training was reported previously with a high risk for lower extremity joint overloading^44^, in support of our results,^45^ Nishiumi et al reported that high impact programs represented by two-legged vertical jumps with jump-rope might improve functional mobility but have no significant change on BMD values of the femoral neck and lumber spine.

When breaking down the effect of each exercise on each joint, the predicted values of joint contact forces during running were highest at the knee, followed by the hip, and then the ankle (load at all joints were higher than walking). Jumping induced a higher load at the knee joint than the hip and ankle joints (load at the knee only was higher than walking), while hopping loaded the ankle joint the most, followed by the knee joint and least at the hip joint (load at the knee and ankle were higher than walking) (Fig. 3). Our study suggests that exercises that improve bone strength are site-specific. Running might enhance bone formation of the whole lower limb joints. Jumping may improve bone formation at the knee joint but, with no or even negative effect on hip and ankle joint regions. Hopping has more effect on the ankle joint than the knee joint, but not the hip joint. Therefore, an osteoporosis exercise program would be better suited to include a combination of these three exercises. However, it is noteworthy that each high-impact exercise was investigated individually, and the combined effects of these exercises were not assessed. Indeed, this is an important factor as a conventional exercise program may be structured to incorporate a variety of high-impact exercises at varying intensities, alongside complementary strength training.^10^

When considering exercise intensity, hip, knee, and ankle joint contact forces increased with higher jumping height but decreased with increased hopping frequency. While there is limited information in the literature regarding the relationship between lower limb joint loading and the intensity level of hopping and jumping exercises, this relation can be partially explained by the amplitude of muscle activation and the cumulative loading on joints during specific exercises. Previous studies have indicated that increased eccentric muscle forces play a role in enhancing vertical jump height.^46^ This is attributed to the correlation between eccentric strength during knee extension and squatting exercises and the resulting jump height. Consequently, an augmentation in jump height might be attributed to increased muscle forces and, subsequently, elevated loading at the joints. On the other hand, increasing hopping frequency is combined with a decrease in hip flexion, knee flexion, and ankle dorsiflexion, which in turn results in an increase in the stiffness of lower limb joints^47,48^ and reduced muscle activations when measured by EMGs.^49,50^ Running showed a different pattern with increasing speed from slow (2.98 m/sec) to moderate (4.25 m/sec) being associated with increased joint contact forces at the hip, knee, and ankle joints. Previous studies have also reported a positive linear relation when assessing running speed (very slow 0.8 m/sec to moderate 3.6 m/sec) with changes in joint contact forces at the hip ^16^ and ankle (4.17m/sec).^51^ However, when running was performed at maximum effort (sprinting at 5.26 m/sec), the increased force at the hip joint was combined with a decreased force at the knee and ankle joints. Large hip muscles, such as the gluteal muscles, become highly active when running fast to counteract the joint moment and to generate higher forces at the hip joint.^52^ In support of our findings, running at a slower speed (2.23 m/sec) was previously reported with increased accumulated loads at the knee compared to faster running (4.38 m/sec).^53^ Peterson et al. explained that the relatively shorter duration of ground contact, combined with the relatively longer distance covered during a loading cycle, reduced the number of loading cycles to traverse an equivalent distance and appeared to mitigate the impact of high loads per stride.

It is worth mentioning that the impact of exercise intensity on the vertical ground reaction force was barely noticeable compared to joint contact forces across all exercises as shown in Table 1. Therefore, the use of ground reaction forces is possibly not a good predictor of peak skeletal loading at the joints and so should not be used alone to assess the intensity of an exercise regimen.^14^ The discrepancies between joint loads and ground reaction forces suggest that other factors, such as muscle activations play an important role in joint loading ^16^. This is related to the fact that muscles exert considerable forces to equilibrate the external moments caused by ground reaction forces during a given motion, serving as protectionary mechanism against damage to the surroundings non-contractile tissue.

The present results should be utilized and interpreted with caution owing to certain limitations. The current study employed static optimization technique. Future studies may investigate joint contact force prediction for high-impact activities using subject-specific MRI-based musculoskeletal models.^54^ Nonetheless, static optimization has been found to adequately replicate muscle activation patterns during walking^38,55^ and hopping^56^, even though the magnitudes of the produced forces remain unverified due to the impracticality of in vivo data acquirement. Another factor is related to the study cohort. Our cohort includes healthy, highly active individuals of both genders, spanning different age groups. Peak joint loading from young adults cannot be generalized to elderly populations^57,58^, similarly for physically non-active to active people, given the differences in motion strategy used to perform an exercise between those groups such as gait length and speed, and standing balance.^59^ Therefore, the current results should not be interpreted for age or gender-specific populations. However, this was beyond the scope of the current study, a follow-up study will address further analysis by generating age- and gender-based subgroups.

The investigation of lower limb joint performance during high-impact activities in this study contributes to the understanding of how such exercise influences joint loading, offering implications for optimizing exercise regimens to enhance bone health. We emphasize that activities like running and hopping, involving weight-bearing and dynamic impacts, appear more favorable for bone formation than jumping alone. Furthermore, our analysis underscores the site-specific nature of exercises in influencing bone strength through unique patterns in joint contact forces under various exercises. Running at varying speeds demonstrated differing impacts on the hip, knee, and ankle joints, while hopping frequency inversely affected joint forces. The current study findings are crucial not only for optimizing exercise regimens to enhance bone health but also for various other aspects, such as designing risk prevention and rehabilitation programs, developing prosthetics, and analyzing sports or occupational activities. Future studies should focus on investigating the stress and strain effects on joints to further understand how joint forces of various exercise types and intensities might trigger bone formation. This can be achieved through coupled musculoskeletal models and finite element models.

## Data Availability

All data produced in the present study are available upon reasonable request to the authors

## 5. Author Contributions

Conceptualization—ZA, BL; Data Curation-ZA, Formal Analysis—ZA, BL, Funding Acquisition— BL, Methodology—ZA, CF, BL, Project Administration—BL, Software—ZA, CF, Supervision—AP, JM, BL, Validation—AP, JM, BL, Visualization—ZA, Writing—Original Draft Preparation—ZA, Writing—Review and Editing-All authors.

