## Supplementary Materials for "Lower limb joint Loading during high-impact activities: implication for bone health"

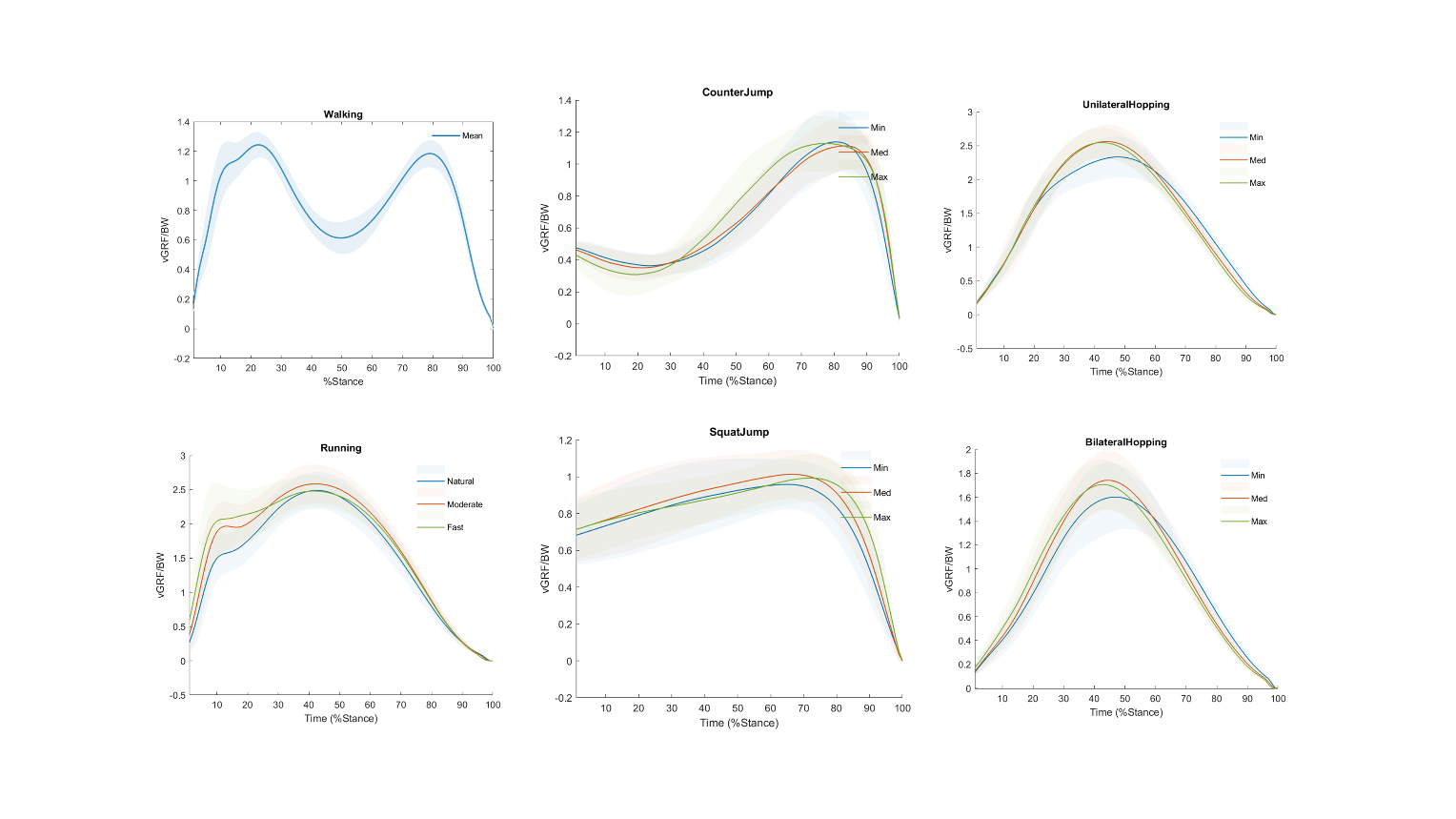

**Supplementary Figure 1.** Mean curves of the vertical ground reaction forces predicted by the musculoskeletal models and normalized by the body weight (vGRF/BW) for all participants. Min, Med, and Max are the effort, minimum, medium, and maximum effort, respectively, for counter movement jump, squat jump, unilateral hopping, and bilateral hopping. Natural, Moderate, and Fast are the minimum, medium, and maximum effort, respectively, for running. Only one level (self-selected speed) was performed by the participate for walking.

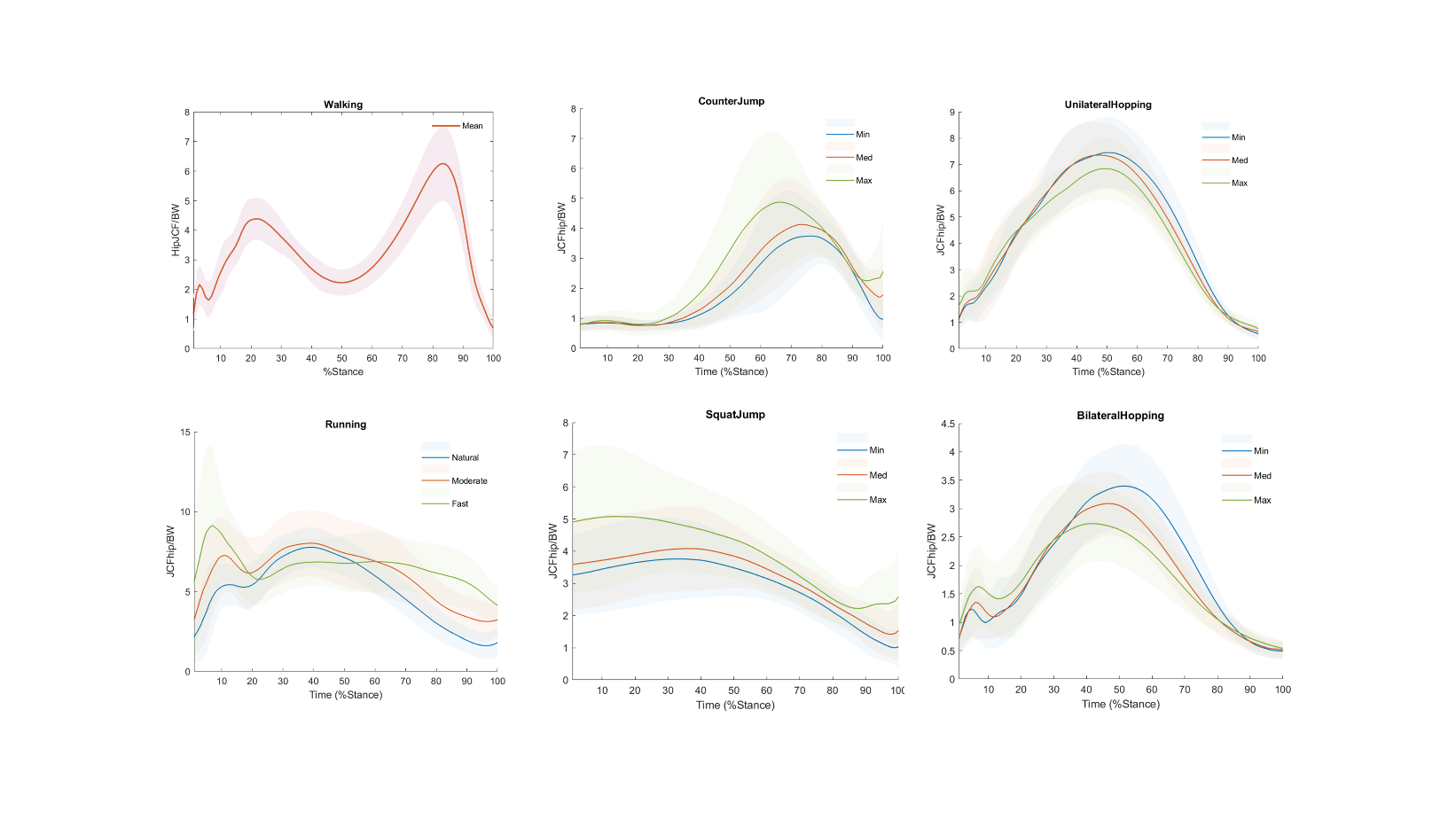

**Supplementary Figure 2.** Mean curves of the resultant hip joint reaction forces predicted by the musculoskeletal models and normalized by the body weight (JCFhip/BW) for all participants. Min, Med, and Max are the effort, minimum, medium, and maximum effort, respectively, for counter movement jump, squat jump, unilateral hopping, and bilateral hopping. Natural, Moderate, and Fast are the minimum, medium, and maximum effort, respectively, for running. Only one level (self-selected speed) was performed by the participate for walking.

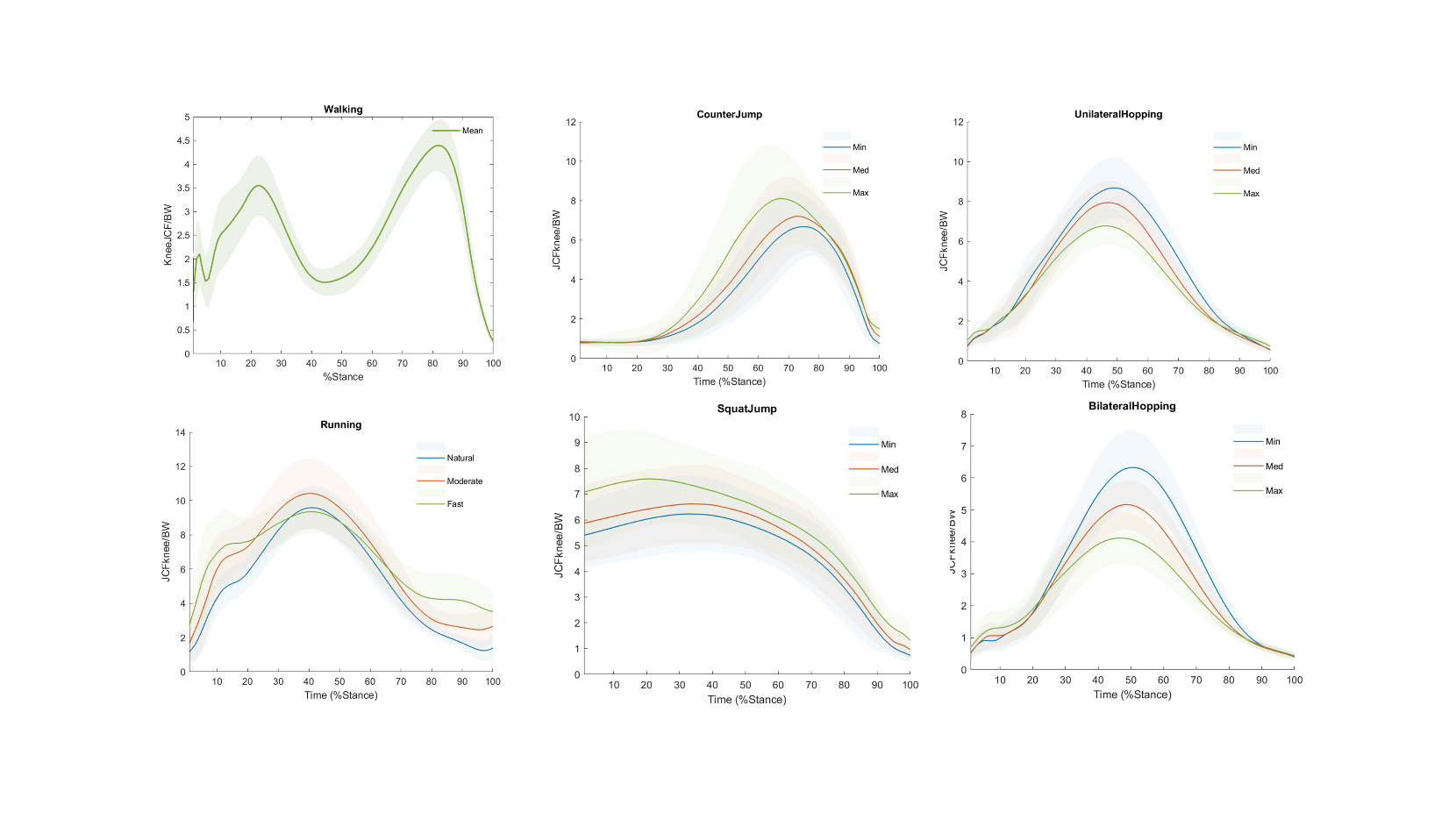

**Supplementary Figure 3.** Mean curves of the resultant knee joint reaction forces predicted by the musculoskeletal models and normalized by the body weight (JCFknee/BW) for all participants. Min, Med, and Max are the effort, minimum, medium, and maximum effort, respectively, for counter movement jump, squat jump, unilateral hopping, and bilateral hopping. Natural, Moderate, and Fast are the minimum, medium, and maximum effort, respectively, for running. Only one level (self-selected speed) was performed by the participate for walking.

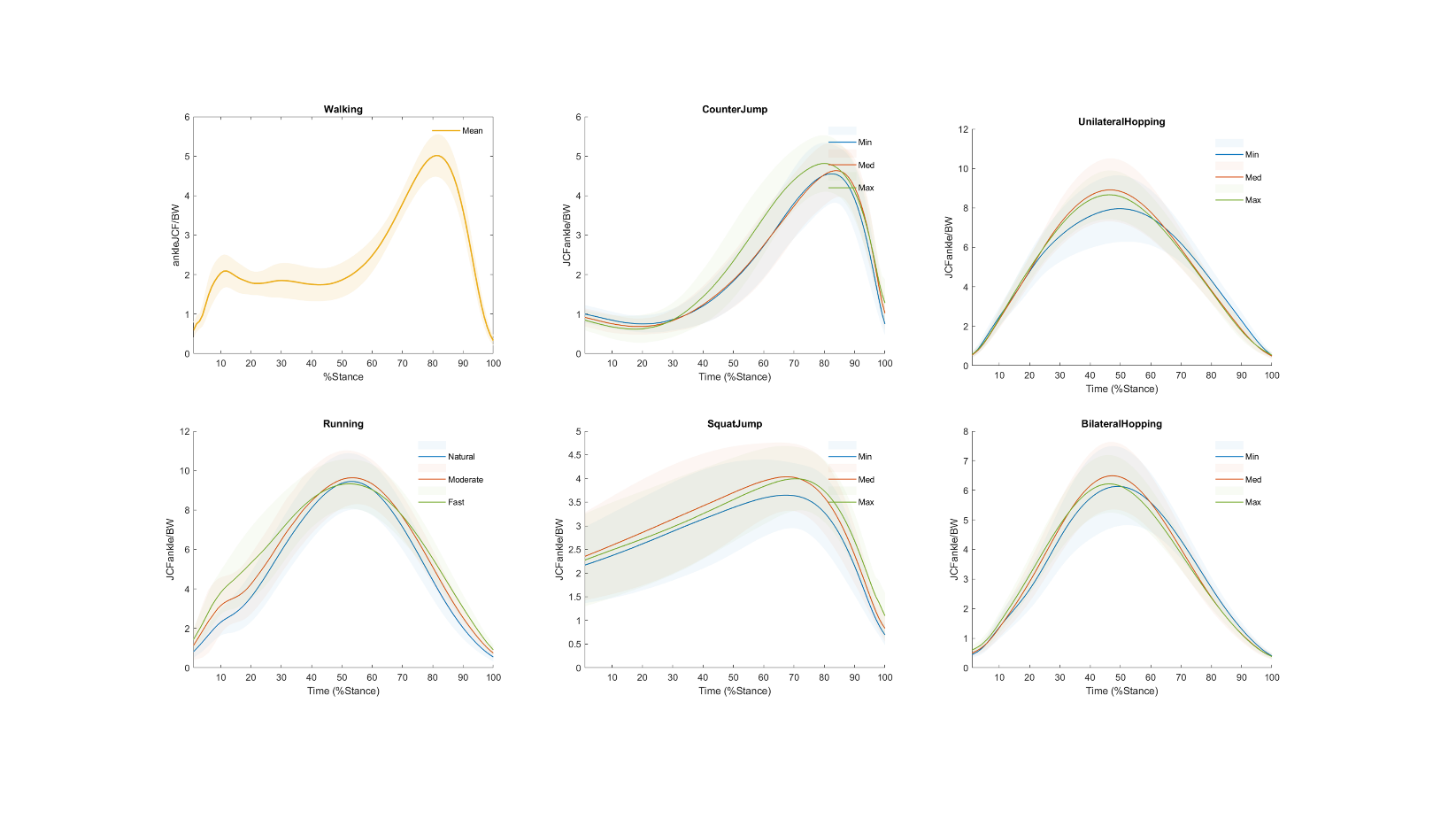

**Supplementary Figure 3.** Mean curves of the resultant ankle joint reaction forces predicted by the musculoskeletal models and normalized by the body weight (JCFankle/BW) for all participants. Min, Med, and Max are the effort, minimum, medium, and maximum effort, respectively, for counter movement jump, squat jump, unilateral hopping, and bilateral hopping. Natural, Moderate, and Fast are the minimum, medium, and maximum effort, respectively, for running. Only one level (self-selected speed) was performed by the participate for walking.

**Table 1.** The estimates, lower/upper limits, and p-values as well as the results from the ANOVA test on the vertical ground reaction force and joint contact force of the hip, knee, and ankle. Exercises with a significant difference (p < .05) compared to walking are marked with an asterisk.

|  | **Exercise** | **Mean Difference (walking-exercise)** | **Std. Error** | **P value** | **95% Confidence Interval for Difference** | |
| --- | --- | --- | --- | --- | --- | --- |
|  |  |  |  |  | **Lower Bound** | **Upper Bound** |
| vGRF | RunningNatural | -1.230^*^ | .043 | .000 | -1.396 | -1.064 |
|  | RunningModerate | -1.337^*^ | .044 | .000 | -1.508 | -1.166 |
|  | RunningFast | -1.338^*^ | .049 | .000 | -1.527 | -1.149 |
|  | SquatJumpsMin | .233^*^ | .029 | .000 | .121 | .346 |
|  | SquatJumpsMed | .200^*^ | .027 | .000 | .096 | .305 |
|  | SquatJumpsMax | .251^*^ | .026 | .000 | .151 | .351 |
|  | CounterJumpsMin | .056 | .030 | 1.000 | -.059 | .171 |
|  | CounterJumpsMed | .096^*^ | .025 | .049 | .000 | .192 |
|  | CounterJumpsMax | .088 | .025 | .111 | -.007 | .184 |
|  | UnilateralHoppingMin | -1.082^*^ | .049 | .000 | -1.273 | -.891 |
|  | UnilateralHoppingMed | -1.312^*^ | .046 | .000 | -1.488 | -1.136 |
|  | UnilateralHoppingMax | -1.308^*^ | .038 | .000 | -1.456 | -1.159 |
|  | BilateralHoppingMin | -.347^*^ | .049 | .000 | -.535 | -.158 |
|  | BilateralHoppingMed | -.483^*^ | .043 | .000 | -.649 | -.318 |
|  | BilateralHoppingMax | -.452^*^ | .035 | .000 | -.586 | -.319 |
| JCFhip | RunningNatural | -1.900* | .240 | .000 | -2.828 | -.972 |
|  | RunningModerate | -3.163* | .400 | .000 | -4.705 | -1.620 |
|  | RunningFast | -5.249* | .652 | .000 | -7.765 | -2.733 |
|  | SquatJumpsMin | 2.250* | .300 | .000 | 1.093 | 3.407 |
|  | SquatJumpsMed | 1.839* | .309 | .000 | .648 | 3.030 |
|  | SquatJumpsMax | .477 | .436 | 1.000 | -1.206 | 2.160 |
|  | CounterJumpsMin | 2.131* | .340 | .000 | .820 | 3.442 |
|  | CounterJumpsMed | 1.715* | .329 | .001 | .447 | 2.984 |
|  | CounterJumpsMax | .680 | .453 | 1.000 | -1.068 | 2.428 |
|  | UnilateralHoppingMin | -1.339* | .281 | .003 | -2.422 | -.255 |
|  | UnilateralHoppingMed | -1.252* | .284 | .010 | -2.348 | -.155 |
|  | UnilateralHoppingMax | -.668 | .238 | .943 | -1.588 | .252 |
|  | BilateralHoppingMin | 2.807* | .201 | .000 | 2.030 | 3.584 |
|  | BilateralHoppingMed | 3.125* | .190 | .000 | 2.393 | 3.858 |
|  | BilateralHoppingMax | 3.375* | .203 | .000 | 2.592 | 4.159 |
| JCFknee | RunningNatural | -5.194* | .225 | .000 | -6.061 | -4.327 |
|  | RunningModerate | -6.034* | .340 | .000 | -7.346 | -4.723 |
|  | RunningFast | -5.460* | .302 | .000 | -6.627 | -4.293 |
|  | SquatJumpsMin | -2.149* | .245 | .000 | -3.093 | -1.204 |
|  | SquatJumpsMed | -2.623* | .263 | .000 | -3.638 | -1.608 |
|  | SquatJumpsMax | -3.811* | .348 | .000 | -5.154 | -2.468 |
|  | CounterJumpsMin | -2.759* | .292 | .000 | -3.885 | -1.634 |
|  | CounterJumpsMed | -3.295* | .307 | .000 | -4.482 | -2.109 |
|  | CounterJumpsMax | -4.370* | .425 | .000 | -6.010 | -2.731 |
|  | UnilateralHoppingMin | -4.237* | .251 | .000 | -5.206 | -3.267 |
|  | UnilateralHoppingMed | -3.519* | .201 | .000 | -4.295 | -2.744 |
|  | UnilateralHoppingMax | -2.334* | .168 | .000 | -2.981 | -1.687 |
|  | BilateralHoppingMin | -1.888* | .196 | .000 | -2.642 | -1.133 |
|  | BilateralHoppingMed | -.703* | .141 | .002 | -1.247 | -.158 |
|  | BilateralHoppingMax | .308 | .142 | 1.000 | -.241 | .858 |
| JCFankle | RunningNatural | -4.491* | .220 | .000 | -5.339 | -3.643 |
|  | RunningModerate | -4.744* | .220 | .000 | -5.591 | -3.897 |
|  | RunningFast | -4.458* | .199 | .000 | -5.224 | -3.692 |
|  | SquatJumpsMin | .939* | .142 | .000 | .390 | 1.488 |
|  | SquatJumpsMed | .605* | .134 | .007 | .088 | 1.121 |
|  | SquatJumpsMax | .729* | .127 | .000 | .238 | 1.220 |
|  | CounterJumpsMin | .130 | .127 | 1.000 | -.361 | .621 |
|  | CounterJumpsMed | .129 | .120 | 1.000 | -.336 | .593 |
|  | CounterJumpsMax | .005 | .118 | 1.000 | -.450 | .460 |
|  | UnilateralHoppingMin | -2.988* | .263 | .000 | -4.001 | -1.975 |
|  | UnilateralHoppingMed | -3.912* | .239 | .000 | -4.833 | -2.990 |
|  | UnilateralHoppingMax | -3.672* | .180 | .000 | -4.366 | -2.978 |
|  | BilateralHoppingMin | -1.171* | .227 | .001 | -2.047 | -.296 |
|  | BilateralHoppingMed | -1.500* | .195 | .000 | -2.254 | -.746 |
|  | BilateralHoppingMax | -1.217* | .156 | .000 | -1.819 | -.616 |
